# *Streptococcus pyogenes* colonization in children aged 24-59 months in The Gambia: Impact of Live Attenuated Influenza Vaccine and associated serological responses

**DOI:** 10.1101/2022.11.27.22282750

**Authors:** Alexander J. Keeley, Danielle Groves, Edwin P. Armitage, Elina Senghore, Ya Jankey Jagne, Hadijatou J. Sallah, Sainabou Drammeh, Adri Angyal, Hailey Hornsby, Gabrielle de Crombrugghe, Pierre Smeesters, Omar Rossi, Martina Carducci, Chikondi Peno, Debby Bogaert, Beate Kampmann, Michael Marks, Helen A. Shaw, Claire E. Turner, Thushan I. de Silva, MRCG Strep A Study Group

## Abstract

**Background:** Immunity to *Streptococcus pyogenes* in high burden settings is poorly understood. We explored *S. pyogenes* nasopharyngeal colonization after intranasal live attenuated influenza vaccine (LAIV) among Gambian children aged 24-59 months, and resulting serological response to 7 antigens.

**Methods:** A post-hoc analysis was performed in 320 children randomized to receive LAIV at baseline (LAIV group) or not (control). *S. pyogenes* colonization was determined by quantitative Polymerase Chain Reaction (qPCR) on nasopharyngeal swabs from baseline (D0), day 7 (D7) and day 21 (D21). Anti-streptococcal IgG was quantified, including a subset with paired serum pre/post *S. pyogenes* acquisition.

**Results:** The point prevalence of *S. pyogenes* colonization ranged from 7-13%. In children negative at D0, *S. pyogenes* was detected at D7 or D21 in 18% of LAIV group and 11% of control group participants (p=0.12). The odds ratio (OR) for colonization over time was significantly increased in the LAIV group (D21 vs D0 OR 3.18, p=0.003) but not in the control group (OR 0.86, p=0.79). The highest IgG increases following asymptomatic colonization were seen for M1 and SpyCEP proteins.

**Conclusions:** Asymptomatic *S. pyogenes* colonization appears modestly increased by LAIV, and may be immunologically significant. LAIV could be used to study influenza-*S. pyogenes* interactions.

## Introduction

*Streptococcus pyogenes* (Group A Streptococcus, Strep A) is responsible for half a million deaths worldwide each year, through severe invasive infections and immune complications including rheumatic heart disease (RHD)[1]. The greatest disease burden is experienced in low- and middle-income countries (LMIC)[1, 2]. Consequently, the World Health Assembly declared in 2018 that a vaccine against *S. pyogenes* is a major global health research priority. An ideal vaccine would substantially reduce *S. pyogenes* transmission, disease, and perhaps colonization, without provoking immune mediated complications [3]. Observational data show a decrease in the incidence of *S. pyogenes* disease with increasing age, possibly due to natural immunity acquired from repeated exposure [4]. Asymptomatic pharyngeal colonization with *S. pyogenes* is recognized, with an estimated point prevalence of 6.6-9.7% in children [4]. Characterizing protective immunity generated by infections and colonization remains a major knowledge gap in designing effective and safe vaccines, particularly from LMIC settings [5-7].

Epidemiological observations have shown an association between invasive *S. pyogenes* infections and respiratory viral infections, especially influenza [8]. *S. pyogenes* was demonstrated to be responsible for substantial mortality in influenza pandemics of 1918 and 2009 [8, 9]. It is important to understand whether respiratory viruses increase colonization with pathogenic bacteria such as *S. pyogenes* in the nasopharynx, as colonization may be a necessary preceding event to pharyngitis and invasive disease. We have recently demonstrated that intranasal live attenuated influenza vaccine (LAIV), used as a surrogate viral challenge agent, induces modest increases in nasopharyngeal *Streptococcus pneumoniae* colonization and density in the 21 days following vaccination in Gambian children [10]. In the current study, we conducted a post-hoc analysis of this randomized controlled trial of LAIV in children aged 24-59 months to explore whether LAIV also induced an increase in nasopharyngeal *S*. *pyogenes* colonization, and whether colonization was associated with a serological response to several *S. pyogenes* antigens.

## Methods

### Study population and design

We carried out a post-hoc observational study nested within a randomized controlled trial of LAIV, studying immunogenicity, viral shedding and microbiome interactions in children aged 24-59 months in The Gambia. Children were randomized 2:1 to receive LAIV at study entry (day 0, D0, LAIV group) or on day 21 (D21, control group), which was the end of active follow up. The study (NCT02972957) was conducted across two years in Sukuta, an urban region in The Gambia, Africa during February to April 2017, and January to March 2018 [10, 11]. Children were recruited to the parent study following community sensitization. All participants were influenza-vaccine naïve and clinically well, with no history of respiratory illness in the prior 14 days. Participants in LAIV and control groups were recruited simultaneously to avoid bias via seasonal variation. Children were immunized with the Northern Hemisphere Russian-backbone Trivalent LAIV (Nasovac-S, Serum Institute of India, Pune, India)[11]. Data on symptoms experienced between D0 and D7 were collected at the D7 visit, and symptoms experienced between D7 and D21 recorded at the D21 visit. Informed consent was sought from parents of all participants, including for subsequent research on their samples. The study was approved by the Joint Medical Research Council/Gambia Government ethics committee (ref: 16193).

### Determination of *S. pyogenes* colonization status

Participants had a nasopharyngeal swab taken at D0 (immediately prior to LAIV receipt), at D7 and D21, using flocked swabs (FLOQSwabs; Copan, USA) stored in RNAprotect (Qiagen, UK). Samples were processed within 4 h of collection and stored at −70°C until further processing. DNA was extracted from 200μl of RNAprotect using the AGOWA Mag Mini DNA extraction kit (LGC Genomics, Berlin, Germany) in combination with phenol-bead beating as previously described[12]. Standard curves were generated via extraction of total genomic DNA from the *S. pyogenes* H293 reference strain, ranging from 1 to 10,000,000 genome copies per µL. Quantitative PCR was performed using QuantStudio™ 5 Real-Time PCR System, using primers and probes to detect the *S. pyogenes* specific gene *speB* (Forward: CTAAACCCTTCAGCTCTTGGTACTG, Reverse: TTGATGCCTACAACAGCACTTTG, Probe: Cy5-CGGCGCAGGCGGCTTCAAC-BHQ2) [13]. A cycle threshold of 40 was defined as positive, but all curves above CT threshold of 35 were checked manually to ensure appearance was consistent with true amplification of the target. Samples were run in triplicate. Where only one of the triplicates showed the presence of *S. pyogenes*, samples were repeated and only defined as positive if at least two triplicates were positive on repeat testing. A participant with positive qPCR result at a given timepoint was considered colonized. All participants were included in the colonization study group. Participants who were negative at baseline were included in the acquisition study group (figure 1).

**Figure 1.**
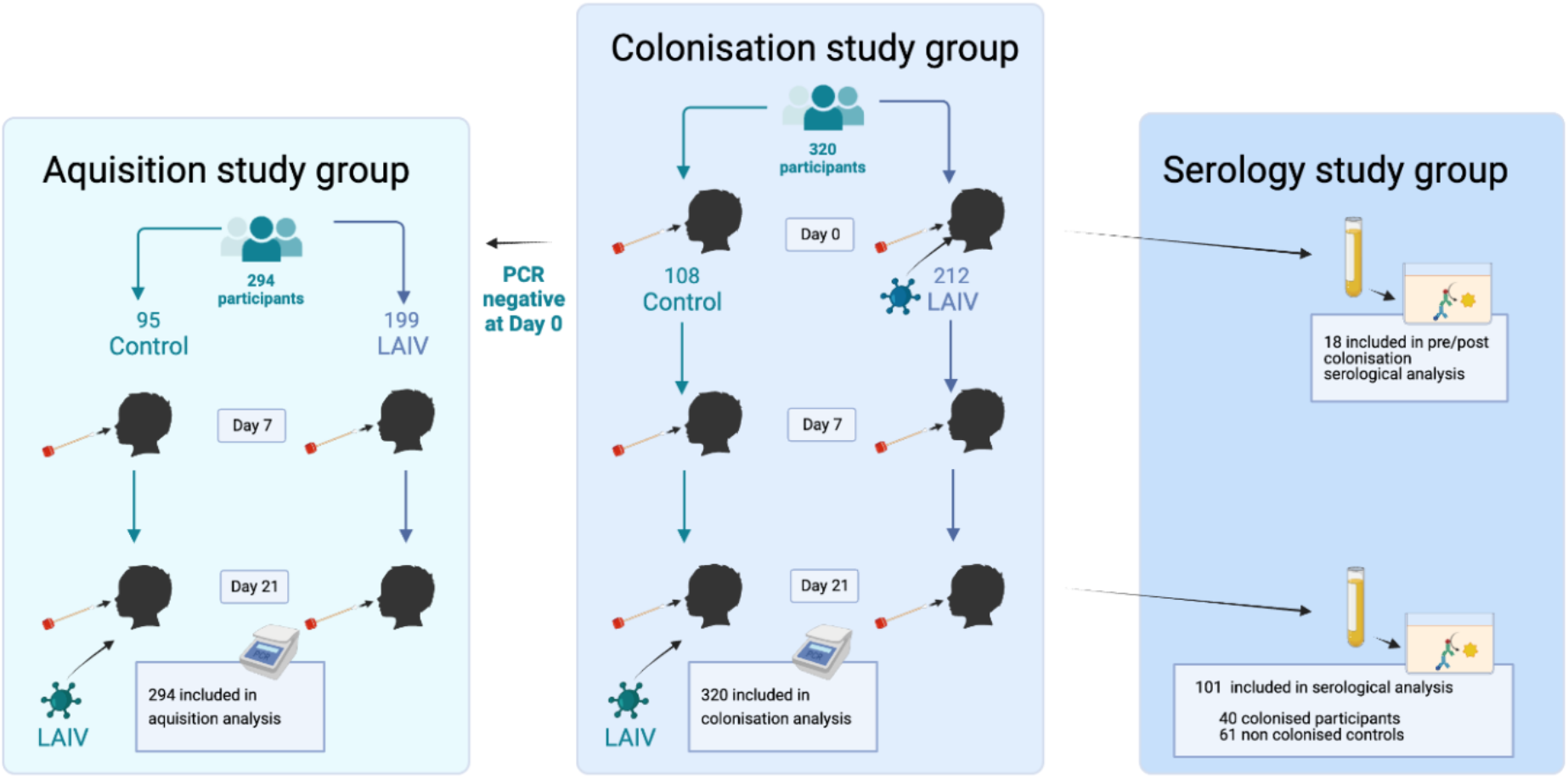
Study profile. 320 participants randomized 2:1 to the LAIV (day 0 vaccine) or control (day 21 vaccine) group. All participants (colonization study group) had S. pyogenes colonization status determined by real-time PCR at day 0, 7 and 21. 294 participants (the acquisition study group) were negative at baseline. Only participants receiving LAIV at day 0 had serum taken at days 0 and 21 (n=212). Serum from day 21 was available from 40/48 participants from LAIV group who were colonized at any time point and from 61/164 randomly selected non-colonized participants. Paired serum (pre/post colonization) was available from 18/35 participants who acquired colonization during the study period. LAIV = Live attenuated Influenza Vaccine. Created with BioRender.com

### Antibody measurement

Blood was collected in serum separation tubes from participants in the LAIV group, for study endpoints on influenza vaccine immunogenicity [11]. For the serology study participants were categorised as colonized if *S. pyogenes* was detected by qPCR at any time point. Participants in the acquisition study group (negative at day 0) were categorised as newly-colonized if *S. pyogenes* was detected at either D7 or D21. All available serum collected on D21 from colonized participants was tested, along with a random selection from non-colonized participants (a 1:1.5 ratio of colonized to non-colonized participants). Of newly-colonized participants, a subgroup of 18 had paired serum (pre/post colonization) available for testing.

Enzyme-linked immunosorbent assay (ELISA) optimization was performed to determine optimal *S. pyogenes* protein coating concentrations, blocking buffer solution, serum dilution and secondary antibody concentration. 96-well flat-bottomed high binding ELISA plates were coated overnight with several recombinant proteins in pH 9.6 carbonate buffer: *S. pyogenes* cell envelope protease (SpyCEP) at 0.16μg/ml, *S. pyogenes* Adhesion and Division protein (SpyAD) at 1μg/ml, M1 protein at 0.5μg/ml, Mac/IdeS protein (henceforth referred to as Mac) at 1 μg/ml, and Collagen binding protein (Cpa) at 0.16 μg/ml. Proteins were provided by National Institute for Biological Standards and Control, MHRA, UK (Supplementary data)[14]. Blocking and serum dilution was performed using 1% casein blocking buffer (ThermoFisher Scientific, USA). Serum was diluted to 1:100 for Cpa and Mac, and 1:800 for M1, SpyCEP and SpyAD, and applied to protein coated wells. Bound IgG was detected with goat anti-human IgG-HRP conjugate (Invitrogen, USA), diluted to 1:500, then incubated with SureBlue TMB 1-Component Peroxidase Substrate (KPL). The reaction was stopped with 1% hydrochloric acid and optical density (OD) was read at 450nm, subtracting background OD. Six washes were performed between each ELISA step with 0.05% Tween-20 in phosphate buffered saline (PBS). In order to quantify anti-protein IgG activity in participant sera, 12 serial three-fold dilutions of pooled human immunoglobulin (Gammanorm, Octapharm) were used to generate standard curves commencing at 1:33.3 dilution (14.85mg/mL) in 1% casein buffer. Seroconversion between D0 and D21 was considered as a 2-fold rise in antibody titre, or 4-fold rise in antibody titre if raw optical density measurements were <0.25 at baseline, as per previously published definitions [15]

In order to measure antibody titres to the additional vaccine antigens Group A carbohydrate (GAC) and Streptolysin O (SLO), an optimized 4-plex (GAC, SLO, SpyCEP, SpyAD) serology assay using the Luminex platform was performed on all available sera using protocol described [16]. All antigens were supplied by GSK Vaccine Institute for Global Health (GVGH). All sera with ELISA data were tested, except two samples from non-colonized participants, where there was no remaining volume. Samples were tested in duplicate at a dilution of 1:8100 alongside standard curves derived from pooled intravenous immunoglobulin (Privigen, CSL Behring). Mean florescence intensity was measured for each antigen with a Luminex 200 instrument (Invitrogen).

### Statistical analysis

All statistical analysis was performed in R (version 4.0.1). Comparison of *S. pyogenes* colonization status and incident acquisition between children who received LAIV and those who did not was performed using chi-squared test. In addition, logistic and generalized mixed effects logistic regression models, accounting for multiple timepoint sampling from individuals, were used to explore the change in colonization status over time within the vaccinated and unvaccinated groups, as previously described [10]. Co-variates included were age in months, sex, the presence of asymptomatic respiratory viruses at baseline, timepoint (D0 vs D7, D0 vs D21) and receipt of LAIV [10, 11].

Samples where optical density measured by ELISA fell below the detection limit of the standard curve, were allocated a random value between zero and the limit of detection. Antibody titres in Luminex data were interpolated using xPonent 4.2 software (Luminex Corporation) using 5-parameter logistic regression. For the 4-plex Luminex assay, where both limit of accurate quantification and detection were characterized, results falling below respective limits were randomly assigned a value between zero and the lowest value of each limit [16]. Antibody data (IVIG–adjusted anti-protein activity in mg/mL for ELISA and Relative Luminex Units (RLU/mL) for Luminex assay) were Log transformed and assessed for normality using QQplot and the shapiro-wilk test. Log transformed antibody quantities between groups were compared using Student’s t-test. Paired Student’s t-test was performed to compare antibody levels before and after *S. pyogenes* colonization from individual participants. A *p* value of <0.05 was considered statistically significant.

## Results

### *S. pyogenes* colonization prevalence and incidence

A total of 320 participants were included in this study, of which 212 received LAIV (Figure 1). Overall, 71/320 (22%) children were colonized with *S. pyogenes* on at least one time point within the 21-day study period. At D0 26/320 (8%) participants were colonized, 13 in the LAIV (6%) and 13 in control (12%) groups (p=0.068, Table S1). In the acquisition study group, 45 (15%) had acquired colonization by D7 or D21 (Table 1), 35/199 (18%) in LAIV group and 10/95 (11%) in control group (Table 1, p=0.12). A logistic regression model was used to explore the odds of new *S. pyogenes* colonization at either D7 or D21 in the acquisition study group (n=294), accounting for age, sex, LAIV receipt and the presence of other respiratory viruses at D0 (Table S2), showing no significant association between LAIV and *S. pyogenes* acquisition (OR 1.92, 0.93-4.31, p=0.09).

**Table 1.** Streptococcus pyogenes acquisition during the 21-day study period. Included below are n=294 in the acquisition study group.

| **Time point** | **N** | **Overall,**<br>**N = 294<sup>1</sup>** | **Control,**<br>**N = 95<sup>1</sup>** | **LAIV,**<br>**N = 199<sup>1</sup>** | **p-value<sup>2</sup>** |
| --- | --- | --- | --- | --- | --- |
| Day 7 | 294 | 18 (6%) | 3 (3%) | 15 (8%) | 0.14 |
| Day 21 | 294 | 35 (12%) | 8 (8%) | 27 (14%) | 0.2 |
| Any time | 294 | 45 (15%) | 10 (11%) | 35 (18%) | 0.12 |

<sup>1</sup>n(%)
<sup>2</sup>Pearson's Chi-squared test

In order to explore changes in colonization status within an individual over time in all children (n=320), a generalized logistic mixed effects regression model was used to explore the odds of *S. pyogenes* colonization over time, accounting for age, sex, LAIV receipt and the presence of other respiratory viruses at D0 (Table S3). As an interaction was observed between LAIV receipt and the D21 timepoint terms in this model, we constructed separate models for the LAIV and control groups. The odds ratio (OR) of *S. pyogenes* colonization was higher at D21 compared to day D0 in the LAIV group (OR 3.18, 95% CI 1.49-6.81, p=0.003, Table 2) but not day 7 (OR 1.30, 95% CI 0.57-2.97, p=0.54). The OR of *S. pyogenes* colonization in the control group was not higher at either D7 (OR 0.34, 95% CI 0.09-1.27, p=0.11) or D21 (OR 0.86, 95% CI 0.3-2.51, p=0.79) when compared to D0 (Table S4).

**Table 2.** Factors associated with S. pyogenes colonization in the live attenuated influenza vaccine (LAIV) group.

|  | OR | 95% CI | p-value |
| --- | --- | --- | --- |
| Day 7 (vs day 0) | 1.30 | 0.57-2.97 | 0.540 |
| Day 21 (vs day 0) | 3.18 | 1.49-6.81 | **0.003** |
| Positive respiratory virus at day 0 | 1.54 | 0.72-3.29 | 0.266 |
| Age in months | 0.97 | 0.93-1.01 | 0.162 |
| Sex Male (vs female) | 0.97 | 0.47-2.04 | 0.945 |

*p*-values for factors associated with *Streptococcus pyogenes* colonization are derived from a generalized logistic mixed effects model, taking into account changes within individuals over time. OR = odds ratio, CI = confidence interval.

Only 17 of 71 (23.9%) participants with *S. pyogenes* detected during the study were colonized at two timepoints, with only 2 (2.8%) colonized at all three study time points (Figure S1). No difference in qPCR quantified *S. pyogenes* density was observed between LAIV and control participants at any timepoint nor over time (Table S5).

We compared symptom data during the study period for all 294 children in the acquisition study group. Ten children had infected skin sores during the 21-day follow up, 9 in the LAIV group and 1 in the control group (5% vs 1%, p=0.2, Table S6). Only three episodes of sore throat were reported, all in the LAIV group (Table S6). Skin sores were more common in those who became colonized (9% vs 2% p=0.05) (Table S6). No statistically significant difference in fever, cough, rhinorrhoea, or sore throat was seen in children who acquired *S. pyogenes* compared to those who did not (Table S6). In the total study group only 6/71 (9%) colonized participants had either a sore throat or an infected skin sore.

### Serological responses to *S. pyogenes* antigens

ELISA-quantified serum antibody titres at D21 in 40 colonized participants were compared with those in 61 randomly selected non-colonized participants (all from the LAIV group due to sera availability). The age was not significantly different between participants included in serological study who were colonized compared to non-colonized controls (median age in months 36 vs 32, p=0.3, Table S6). Sera from colonized participants demonstrated significantly higher IVIG-adjusted IgG levels to M1, SpyCEP, SpyAD and Mac, but not Cpa, compared to non-colonized participants (Figure 2A). Paired serum was available for 18/35 newly-colonized participants. Mean M1- and SpyCEP-specific IgG titres were significantly increased at D21 compared to D0, but not IgG titers to SpyAD, Mac or Cpa (Figure 2B). The proportion of newly-colonized participants who seroconverted pre/post colonization was greatest for M1 and SpyCEP (Figure 2C). To explore serological responses further, a recently described 4-plex assay was used to quantify antibodies to additional *S. pyogenes* vaccine antigens GAC and SLO, along with SpyCEP and SpyAD. This assay was tested on 99/101 samples with ELISA-quantified anti-*S. pyogenes* titres [16]. Sera from colonized children demonstrated significantly higher titres to GAC, SLO and SpyCEP but not to SpyAD (Figure 3A). In the subset of participants with *S. pyogenes* acquisition during the 21-day study period (n=18), differences in IgG titers at D0 and D21 were not statistically significant (Figure 3B). Antibody titres to SpyCEP and SpyAD that were measured by both techniques were well correlated (Figure S2). Removing the participants with sore throat or infected skin sores and colonization during the study had no significant impact on the serological results, nor did excluding participants colonized on D21 only, nor applying a more conservative definition of seroconversion (4-fold increase in IgG titre regardless of baseline optical density) (Figures S3, S4, S5).

**Figure 2.**
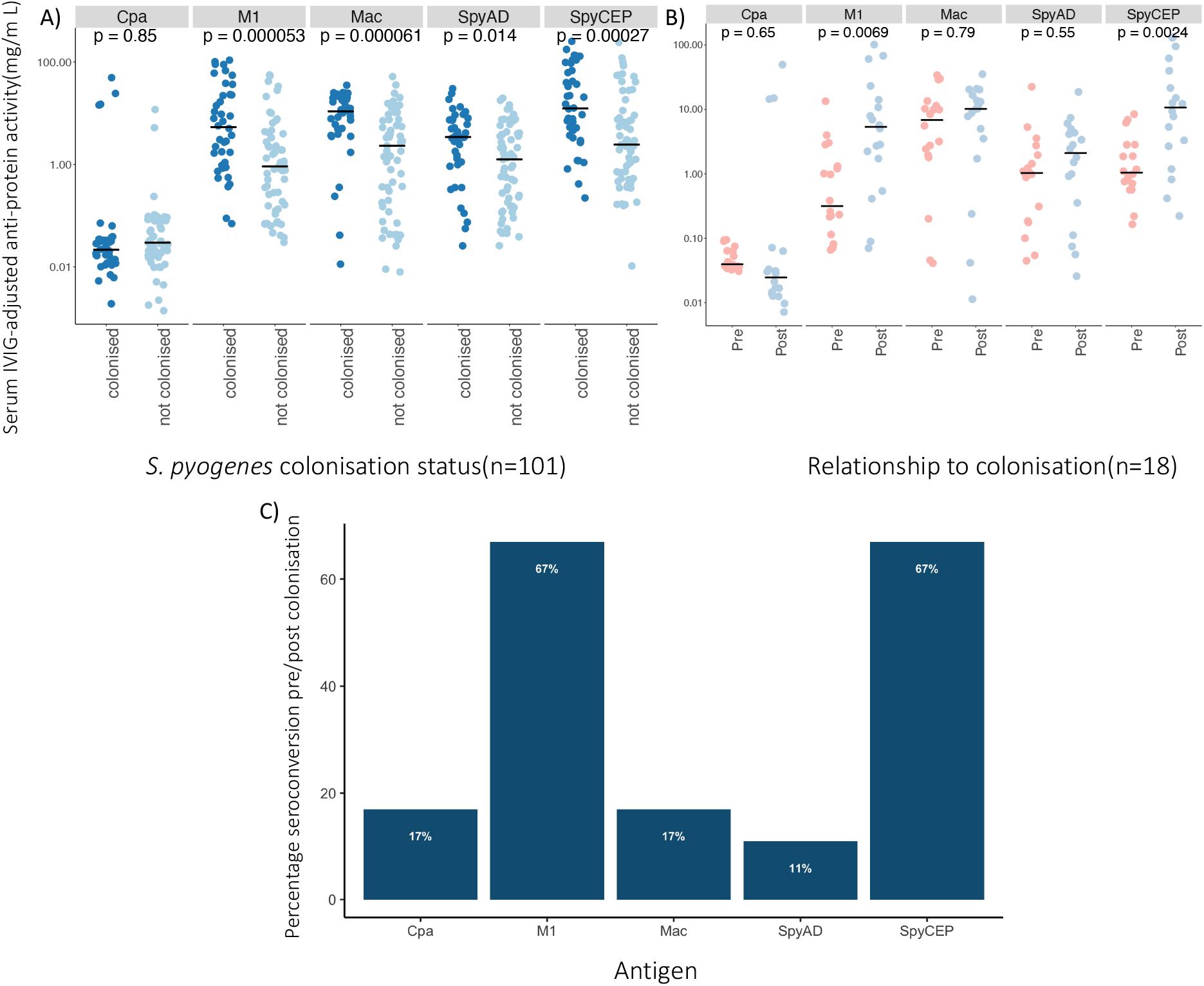
Serological responses to S. pyogenes colonization measured by ELISA. **A**. Comparison of anti-protein IgG activity to Cpa, M1, Mac, SpyCEP, and SpyAD in participants (n=101) according to anytime S. pyogenes colonization status. **B**. Paired comparison of anti-protein IgG activity to Cpa, M1, Mac, SpyCEP, and SpyAD between day 0 and day 21 in newly colonized participants (n=18). Log_10_ transformed IVIG-adjusted anti-protein activity was compared with t-tests (unpaired and paired respectively), horizontal line depicts the median value. **C**. Percentage of study participants (n=18) acquiring S. pyogenes during the study who seroconverted between day 0 and day 21.

**Figure 3.**
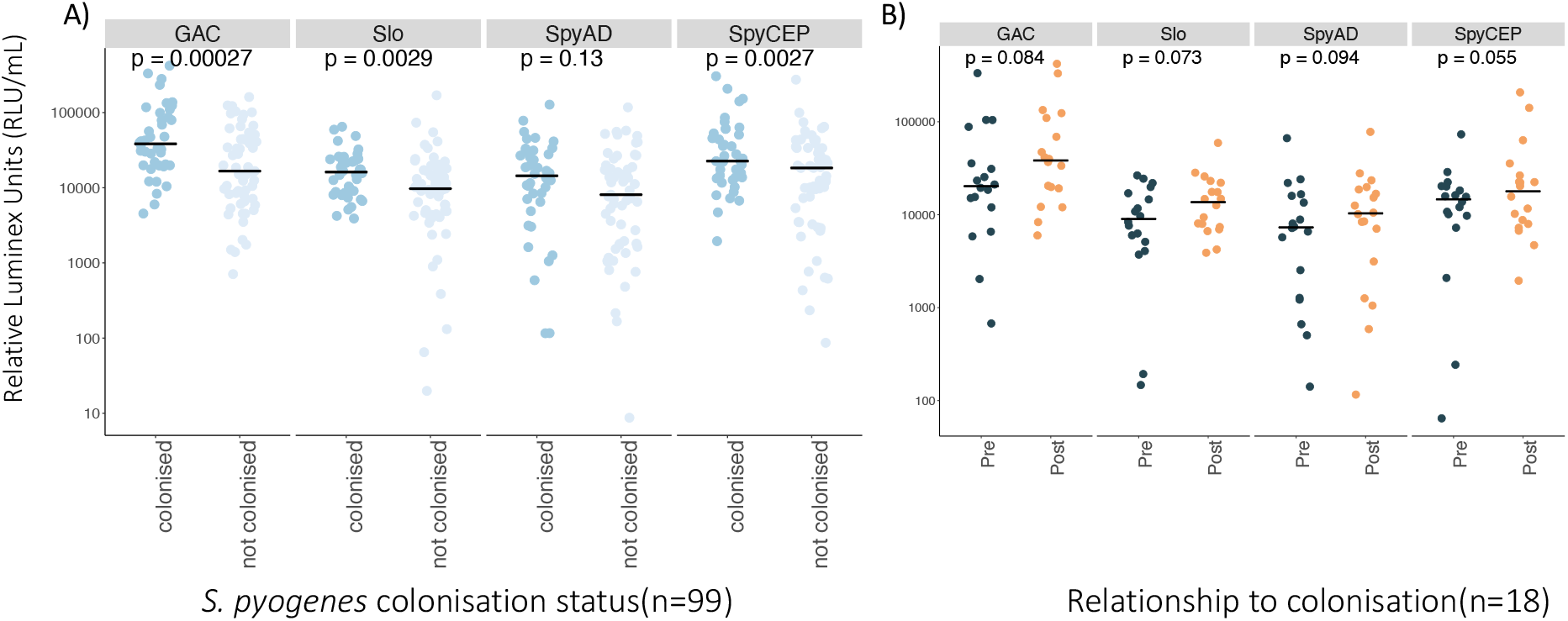
Serological responses to S. pyogenes colonization measured by Luminex 4-plex. **A**. Comparison of IgG activity to GAC, SLO, SpyCEP, and SpyAD in participants (n=99) according to anytime S. pyogenes colonization status. **B**. Paired comparison of IgG activity to GAC, SLO, SpyCEP, and SpyAD between D0 and D21 in newly-colonized participants (n=18). Log_10_ transformed IVIG-adjusted anti-antigen activity was compared with t-tests (unpaired and paired respectively). Horizontal bar depicts the median value.

## Discussion

In this post-hoc study of a randomized controlled trial set in The Gambia, we report to our knowledge the first analysis examining the impact of LAIV on *S. pyogenes* nasopharyngeal colonization in children. Using PCR to define colonization events, we observed a prevalence of 8% at the start of the study in children aged 24-59 months. Our data demonstrate only a modest impact of LAIV on *S. pyogenes* colonization rates. We observed non-significantly increased rates and odds of new colonization in the LAIV group and increased odds of colonization at D21 compared to baseline in the LAIV group only. During the short 21-day study duration, 22% of children were colonized at one or more timepoints, with most children (73%) only colonized at one timepoint. The dynamic picture of colonization we observed may be partially influenced by LAIV administration. It may also reflect how commonly children are exposed to *S. pyogene*s in areas with a high burden of *S. pyogenes* disease and our findings are broadly in keeping with published estimates from LMICs [4, 17].

*S. pyogenes* was demonstrated to be responsible for substantial mortality in influenza pandemics of 1918 and 2009 [8, 9]. While both epidemiological and mechanistic observations demonstrate that respiratory viruses, including influenza, are associated with severe *S. pyogenes* disease, their role in modulating the host-pathogen interaction is complex and poorly understood, particularly in relation to colonization of the pharynx [8, 9, 18-21]. As respiratory viruses frequently infect the colonizing sites of bacteria, it is important to understand their impact on potentially pathogenic bacteria such as *S. pyogenes*. While mice administered a non-lethal challenge with *S. pyogenes* following an influenza challenge frequently had a severe and fatal disease course [21], mice vaccinated with LAIV were protected from *S. pyogenes* superinfection [20], but few models have specifically investigated the impact of respiratory viruses on pharyngeal colonization [22]. With the recent establishment of a *S. pyogenes* human challenge model, co-challenge studies with LAIV and *S. pyogenes* could reveal mechanistic insights into the interaction between influenza virus, *S. pyogenes*, and host immune responses [23]. A co-challenge study with *S. pneumoniae* and LAIV has shown how virus-induced inflammatory responses and impaired innate immune responses promote bacterial colonization [24]. The data provide further evidence that the impact of LAIV on colonization with potentially pathogenic bacteria is only modest and supports the wider rollout of LAIV to reduce influenza disease and its complications in LMICs.

We also demonstrate that asymptomatic *S. pyogenes* colonization leads to seroconversion to several antigens representative of different stages in *S. pyogenes* infection, with differences in antibody magnitude across the different targets measured[14]. Higher serological responses to *S. pyogenes* antigens were observed in children colonized at any timepoint in our study compared to non-colonized controls. Given that prior exposure to *S. pyogenes* in all children in this cohort may be similar, these differences likely reflect recent exposure. The most notable responses were seen to full length streptococcal M1 protein and to the envelope protease SpyCEP when measured by ELISA, including a rise in titres in paired sera taken from before and after colonization. Reactivity to full length M1 protein likely reflects activity to conserved M protein region rather than type-specific reactivity, given that *emm*1 isolates have not been identified in The Gambia[25, 26]. IgG antibodies to other antigens, SpyAD, Mac, GAC and SLO, were higher in children where *S. pyogenes* colonization had been detected, but not Cpa. Noting that the antigens tested via ELISA and Luminex were obtained from different sources, there was broad concurrence between the two serological assays, except that IgG to SpyAD was significantly higher in colonized compared to non-colonized participants when measured by ELISA and not by Luminex. Nonetheless, our findings suggest that asymptomatic pharyngeal colonization with *S. pyogenes* may indeed elicit an IgG immunological response to multiple antigens.

Whilst “true” *S. pyogenes* colonization has been defined as colonization *without* a serological response, the definition risks simplification of a complex and dynamic state influenced by host-immunity, bacterial characteristics, and environmental factors. This serologically inactive phenotype has been described with minimal serum antibody response to SLO and anti-DNAase B (7, 32). Our data shows that serological responses are variable following asymptomatic colonization to different proteins. Furthermore, in practice colonization is often defined as an asymptomatic person with detectable *S. pyogenes* without serial serological testing. In detailed longitudinal analyses, seroconversion has been documented following asymptomatic acquisition of *S. pyogenes* in the USA and Egypt, with highest proportion of seroconversions to type-specific M peptides and SpyAD [15, 27]. In another cohort study in the USA, seroconversion to type-specific M protein frequently occurred following asymptomatic acquisition and conferred protection from homologous strain reinfection [28].

Protection following asymptomatic colonization and historical *S. pyogenes* intranasal vaccination has been observed, but the responsible mechanisms and serological corelates of protection have not been identified [29-34]. Early prospective observation demonstrated the emergence of acute rheumatic fever following asymptomatic *S. pyogenes* infection [35]. Furthermore, emerging data show that patients with rheumatic fever have serological activity to significantly more *S. pyogenes* M peptides and conserved antigens compared to matched controls, including activity following asymptomatic pharyngeal colonization [36, 37]. It is therefore possible that asymptomatic colonization could play a role in the pathological immunity that leads to acute rheumatic fever in endemic settings and warrants further exploration.

Our study has several key limitations. Firstly, it was a post-hoc analysis of a study principally powered for assessing the impact of LAIV on *S. pneumoniae* colonization density, and interactions between the host microbiome and LAIV-specific immunity. The study was not powered to detect any impact of LAIV on *S. pyogenes* density. LAIV is used as a proxy for natural influenza infection and may not reflect the true dynamics of *S. pyogenes* colonization during natural influenza infection. We defined colonization events by PCR and not microbiological culture of *S. pyogenes*, which is considered the gold standard for defining the presence of *S. pyogenes*. We have previously demonstrated that the point prevalence of pyoderma in children under 5 from the same community is high at 17.4%, with *S. pyogenes* cultured from 51% of pyoderma lesions [38]. We did not perform microbiological culture on infected skin lesions or from sore throats in this study to be able to definitively categorize these episodes as symptomatic S. pyogenes events. Nonetheless the exclusion of all participants with either a sore throat or skin sore did not dramatically alter the serological findings. Due to the availability of serum samples, we only measured *S. pyogenes-*specific antibodies in in the LAIV group, so no comparison between vaccinated and unvaccinated groups was possible. Finally, PCR to a single preserved and specific *S. pyogenes* target does not allow for assessment of *emm*-type specific immunity, which is an historically important consideration for *S. pyogenes* serological activity.

Nonetheless, our study does provide several important findings. Understanding both naturally occurring protective immunity and pathological autoimmunity to *S. pyogenes* from settings with the highest disease burden is of paramount importance to progress towards a safe and effective *S. pyogenes* vaccine. Our study adds further evidence that asymptomatic colonization may be immunologically significant, particularly in the context of influenza co-infection. Further research combining longitudinal observation, with detailed microbiological and immunological investigation should be prioritised from areas of high disease prevalence to gain a deeper understanding of immunity to this major human pathogen.

## Data Availability

All data produced in the present study are available upon reasonable request to the authors

## Acknowledgements

The authors would like to acknowledge and thank the Strep A Study Group at Medical Research Council (MRC) Unit The Gambia at London School of Hygiene & Tropical Medicine (LSHTM), Sona Jabang, Annette Erhart, Saffiatou Darboe, Abdul Karim Sesay, Peggy-Estelle Tiencheu, Saikou Y. Bah, Lamin Jaiteh, Karen Forrest, Anna Roca, Fatoumata Camara; Peggy-Estelle Tiencheu, Aru-Kumba Baldeh, Grant Mackenzie, Martin Antonio; GSK Vaccine Institute of Global Health, particularly Danilo Moriel and Luisa Masai for the technical support and provision of SpyCEP, Slo, GAC and SpyAD antigens for measuring IgG in Luminex 4-plex assay; the dedicated team of field and nursing staff, especially Sulayman Bah and Janko Camara, who delivered the study at MRC Unit The Gambia at LSHTM; Isatou Ndow for clinical trial organization; the research support and clinical trials support offices at the Unit the Serum Institute of India for donating the vaccines for the study. Most importantly we thank the study participants and their parents who took part in the study.

## Footnote Page

### Funding

This research was funded by Wellcome Trust in part by an Intermediate Clinical Fellowship award to TIdS [Grant no: 110058/Z/15/Z] and in part by a Clinical PhD fellowship in Global Health award to AJK [Grant no: 225467/Z/22/Z]. For the purpose of open access, the author has applied a CC BY public copyright licence to any Author Accepted Manuscript version arising from this submission.

### Previous presentation

The preliminary data from this study were presented at Lancefield Symposium for Streptococcal and Streptococcal Diseases, Stockholm 2022, and at European Congress of Clinical Microbiology and Infectious Diseases 2021, Online.

