## Supplementary material for "*Streptococcus pyogenes* colonization in children aged 24-59 months in The Gambia: Impact of Live Attenuated Influenza Vaccine and associated serological responses"

### Supplementary data

#### 1. Protein sequences for Streptococcus pyogenes ELISA

**> M1 48 kDa**
MGNGDGNPREVIEDLAANNPAIQNIRLRYENKDLKARLENAMEVAGRDFKRAEELEKAKQ
ALEDQRKDLETKLKELQQDYDLAKESTSWDRQRLEKELEEKKEALELAIDQASRDYHRAT
ALEKELEEKKKALELAIDQASQDYNRANVLEKELETITREQEINRNLLGNAKLELDQLSS
EKEQLTIEKAKLEEEKQISDASRQSLRRDLDASREAKKQVEKDLANLTAELDKVKEDKQI
SDASRQGLRRDLDASREAKKQVEKDLANLTAELDKVKEEKQISDASRQGLRRDLDASREA
KKQVEKALEEANSKLAALEKLNKELEESKKLTEKEKAELQAKLEAEAKALKEQLAKQAEE
LAKLRAGKASDSQTPDTKPGNKAVPGKGQAPQAGTKPNQNKAPMKETKRQLEHHHHHH

**>SpyCEP 61 kDa**
MGDELSTMSEPTITNHAQQQAQHLTNTELSSAESKSQDTSQITLKTNREKEQSQDLVSEP
TTTELADTDAASMANTGSDATQKSASLPPVNTDVHDWVKTKGAWDKGYKGQGKVVAVIDT
GIDPAHQSMRISDVSTAKVKSKEDMLARQKAAGINYGSWINDKVVFAHNYVENSDNIKEN
QFEDFDEDWENFEFDAEAEPKAIKKHKIYRPQSTQAPKETVIKTEETDGSHDIDWTQTDD
DTKYESHGMHVTGIVAGNSKEAAATGERFLGIAPEAQVMFMRVFANDIMGSAESLFIKAI
EDAVALGADVINLSLGTANGAQLSGSKPLMEAIEKAKKAGVSVVVAAGNERVYGSDHDDP
LATNPDYGLVGSPSTGRTPTSVAAINSKWVIQRLMTVKELENRADLNHGKAIYSESVDFK
DIKDSLGYDKSHQFAYVKESTDAGYNAQDVKGKIALIERDPNKTYDEMIALAKKHGALGV
LIFNNKPGQSNRSMRLTANGMGIPSAFISHEFGKAMSQLNGNGTGSLEFDSVVSKAPSQK
GNEMNHFSNWGLTSDLEHHHHHH

**>Cpa 77 kDa**
MGKTVFGLVESSTPNAINPDSSSEYRWYGYESYVRGHPYYKQFRVAHDLRVNLEGSRSYQ
VYCFNLKKAFPLGSDSSVKKWYKKHDGISTKFEDYAMSPRITGDELNQKLRAVMYNGHPQ
NANGIMEGLEPLNAIRVTQEAVWYYSDNAPISNPDESFKRESESNLVSTSQLSLMRQALK
QLIDPNLATKMPKQVPDDFQLSIFESEDKGDKYNKGYQNLLSGGLVPTKPPTPGDPPMPP
NQPQTTSVLIRKYAIGDYSKLLEGATLQLTGDNVNSFQARVFSSNDIGERIELSDGTYTL
TELNSPAGYSIAEPITFKVEAGKVYTIIDGKQIENPNKEIVEPYSVEAYNDFEEFSVLTT
QNYAKFYYAKNKNGSSQVVYCFNADLKSPPDSEDGGKTMTPDFTTGEVKYTHIAGRDLFK
YTVKPRDTDPDTFLKHIKKVIEKGYREKGQAIEYSGLTETQLRAATQLAIYYFTDSAELD
KDKLKDYHGFGDMNDSTLAVAKILVEYAQDSNPPQLTDLDFFIPNNNKYQSLIGTQWHPE
DLVDIIRMEDKKEVIPVTHNLTLRKTVTGLAGDRTKDFHFEIELKNNKQELLSQTVKTDK
TNLEFKDGKATINLKHGESLTLQGLPEGYSYLVKETDSEGYKVKVNSQEVANATVSKTGI
TSDETLAFENNKEPLEHHHHHH

**>Mac 36 kDa**
MGSANQEIRYSEVTPYHVTSVWTKGVTPPANFTQGEDVFHAPYVANQGWYDITKTFNGKD
DLLCGAATAGNMLHWWFDQNKDQIKRYLEEHPEKQKINFNGEQMFDVKEAIDTKNHQLDS
KLFEYFKEKAFPYLSTKHLGVFPDHVIDMFINGYRLSLTNHGPTPVKEGSKDPRGGIFDA
VFTRGDQSKLLTSRHDFKEKNLKEISDLIKKELTEGKALGLSHTYANVRINHVINLWGAD
FDSNGNLKAIYVTDSDSNASIGMKKYFVGVNSAGKVAISAKEIKEDNIGAQVLGLFTLST
GQDSWNQTNLEHHHHHH

**>SpyAD 89 kDa**
MGDRASGETKASNTHDDSLPKPETIQEAKATIDAVEKTLSQQKAELTELATALTKTTAEINH
LKEQQDNEQKALTSAQEIYTNTLASSEETLLAQGAEHQRELTATETELHNAQADQHSKET
ALSEQKASISAETTRAQDLVEQVKTSEQNIAKLNAMISNPDAITKAAQTANDNTKALSSE

LEKAKADLENQKAKVKKQLTEELAAQKAALAEKEAELSRLKSSAPSTQDSIVGNNTMKAP
QGYPLEELKKLEASGYIGSASYNNYYKEHADQIIAKASPGNQLNQYQDIPADRNRFVDPD
NLTPEVQNELAQFAAHMINSVRRQLGLPPVTVTAGSQEFARLLSTSYKKTHGNTRPSFVY
GQPGVSGHYGVGPHDKTIIEDSAGASGLIRNDDNMYENIGAFNDVHTVNGIKRGIYDSIK
YMLFTDHLHGNTYGHAINFLRVDKHNPNAPVYLGFSTSNVGSLNEHFVMFPESNIANHQR
FNKTPIKAVGSTKDYAQRVGTVSDTIAAIKGKVSSLENRLSAIHQEADIMAAQAKVSQLQ
GKLASTLKQSDSLNLQVRQLNDTKGSLRTELLAAKAKQAQLEATRDQSLAKLASLKAALH
QTEALAEQAAARVTALVAKKAHLQYLRDFKLNPNRLQVIRERIDNTKQDLAKTTSSLLNA
QEALAALQAKQSSLEATIATTEHQLTLLKTLANEKEYRHLDEDIATVPDLQVAPPLTGVK
PLSYSKIDTTPLVQEMVKETKQLLEASARLAAENTSLVAEALVGQTSEMVASNAIVSKIT
SSITQPSSKTSYGSGSSTTSNLISDVDESTQRLEHHHHHH

**Supplementary Figures and Tables**

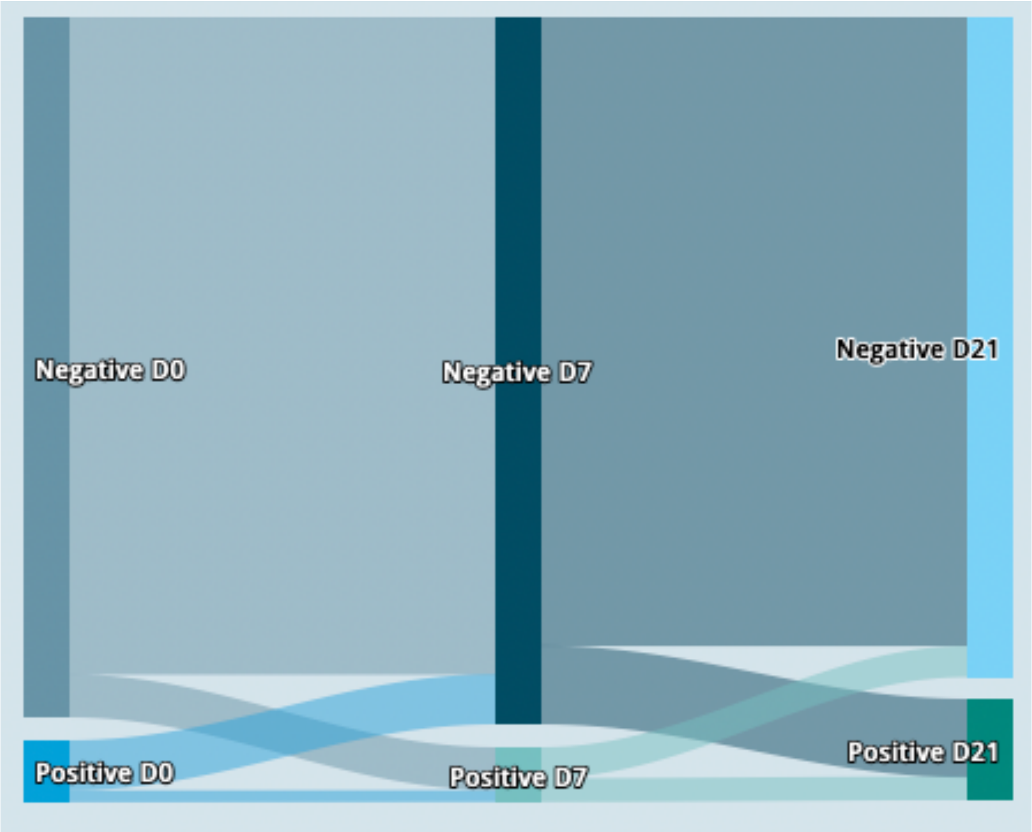


**Figure S1: Colonization status over time.** Sankey plot demonstrating the proportion of participants with positive and negative colonization status at each time point in the study. Colonization status determined by quantitative PCR targeting *SpeB*.

**Table S1. *S. pyogenes* colonization during the study period.** Colonization was determined by a *SpeB* real-time polymerase chain reaction at day 0, day 7 and day 21 of the study in children in the LAIV and control groups.

| **Time point** | **N** | **Overall**, N = 320[1] | **LAIV**, N = 212[1] | **Control**, N = 108[1] | **p-value**[2] |
|---|---|---|---|---|---|
| **Baseline** | 320 | | | | 0.068 |
| negative | | 294 (92%) | 199 (94%) | 95 (88%) | |
| positive | | 26 (8.1%) | 13 (6.1%) | 13 (12%) | |
| **Day 7** | 320 | | | | 0.7 |
| negative | | 297 (93%) | 196 (92%) | 101 (94%) | |
| positive | | 23 (7.2%) | 16 (7.5%) | 7 (6.5%) | |
| **Day 21** | 320 | | | | 0.4 |
| negative | | 277 (87%) | 181 (85%) | 96 (89%) | |
| positive | | 43 (13%) | 31 (15%) | 12 (11%) | |
| **Anytime** | 320 | | | | 0.8 |
| negative | | 249 (78%) | 164 (77%) | 85 (79%) | |
| positive | | 71 (22%) | 48 (23%) | 23 (21%) | |

[1] n (%)

[2] Pearson's Chi-squared test

**Table S2. Factors associated with new colonization at D7 or D21 in the acquisition study group (n=294)**

| Characteristic | OR[1] | 95% CI[1] | p-value |
|---|---|---|---|
| Age in months | 0.99 | 0.96, 1.03 | 0.7 |
| Intervention (Vaccinated) | 1.92 | 0.93, 4.31 | 0.094 |
| sex | | | |
| F | — | — | |
| M | 1.34 | 0.69, 2.62 | 0.4 |
| Positive respiratory virus at day 0 | 1.71 | 0.88, 3.28 | 0.11 |

[1] OR = Odds Ratio, CI = Confidence Interval

*p values for factors associated with new *Streptococcus pyogenes* acquisition within the acquisition study group (n=294) are derived from a logistic regression model with a composite outcome of colonization at either D7 or D21.

**Table S3 Factors associated with *S. pyogenes* colonization prevalence in the entire cohort.**

| | *OR* | *95% CI* | *p-value*[1] |
|---|---|---|---|
| *Intervention (vaccinated)* | 0.41 | 0.14-1.14 | 0.09 |
| *Day 21 (vs day 0)* | 0.88 | 0.34-2.28 | 0.8 |
| *Day 7 (vs day 0)* | 0.41 | 0.14-1.22 | 0.11 |
| *Positive respiratory virus at day 0* | 1.79 | 0.9-3.54 | 0.09 |
| *Age* | 0.98 | 0.94-1.02 | 0.29 |
| *Sex Male (vs female)* | 0.97 | 0.5-1.88 | 0.93 |
| *Intervention (vaccinated): Day 21* | 3.86 | 1.12-13.27 | **0.03** |
| *Intervention (vaccinated): Day 7* | 3.2 | 0.8-12.77 | 0.1 |

[1]p values for factors associated with Streptococcus pyogenes colonization are derived from a generalised logistic mixed-effect model, taking into account changes within individuals over time. Given the significant interaction between Intervention and Visit variables, the model was tested on both the vaccinated (intervention) and unvaccinated groups.

**Table S4. Factors associated with *S. pyogenes* colonization prevalence in the unvaccinated group.**

| | *OR* | *95% CI* | *p-value*[1] |
|---|---|---|---|
| *Day 21 (vs day 0)* | 0.86 | 0.3-2.51 | 0.79 |
| *Day 7 (vs day 0)* | 0.34 | 0.09-1.27 | 0.11 |
| *Positive respiratory virus at day 0* | 2.55 | 0.53-12.34 | 0.25 |
| *Age* | 1 | 0.92-1.1 | 0.95 |
| *Sex Male (vs female)* | 0.98 | 0.2-4.73 | 0.98 |

[1]p values for factors associated with *Streptococcus pyogenes* colonization in the live attenuated influenza vaccine group are derived from a generalised logistic mixed-effect model, taking into account changes within individuals over time.

**Table S5. Comparison of *S. pyogenes* density (log10 cpoies /mL) measured by qPCR in colonized participants (n=71) between vaccinated and unvaccinated groups at each time point of the study**

| | Study group | | |
|---|---|---|---|
| | LAIV[1] | Control[1] | p-value[2] |
| D0 | 3.80 (3.49, 4.35) | 3.70 (3.24, 4.24) | 0.6 |
| D7 | 3.99 (3.73, 4.97) | 4.60 (4.20, 6.43) | 0.2 |
| D21 | 3.70 (3.40, 4.25) | 4.34 (3.49, 5.41) | 0.12 |

[1] Median (IQR)
[2] Welch Two Sample t-test

**Table S6. Comparison of symptoms reported by study participants by day 21 of the study in those who did or did not acquire *S. pyogenes* colonization.**

| | Acquisition status | | |
|---|---|---|---|
| **Sypmtoms** | **No**, N = 249[1] | **Yes**, N = 45[1] | **p-value**[2] |
| fever | 51 (20%) | 12 (27%) | 0.4 |
| cough | 94 (38%) | 14 (31%) | 0.4 |
| runny nose | 118 (47%) | 26 (58%) | 0.2 |
| sore throat | 1 (0.4%) | 2 (4.4%) | 0.062 |
| skin sore | 6 (2.4%) | 4 (8.9%) | 0.050 |

[1] n (%)

[2] Pearson's Chi-squared test; Fisher's exact test

**Table S7. Comparison of age (in months) between colonized and non-colonized participants included in serological study (n=101).**

| Characteristic | colonised, N = 40[1] | non-colonised, N = 61[1] | p-value[2] |
|---|---|---|---|
| age | 36 (29, 43) | 32 (28, 39) | 0.3 |

[1] Median (IQR)
[2] Wilcoxon rank sum test

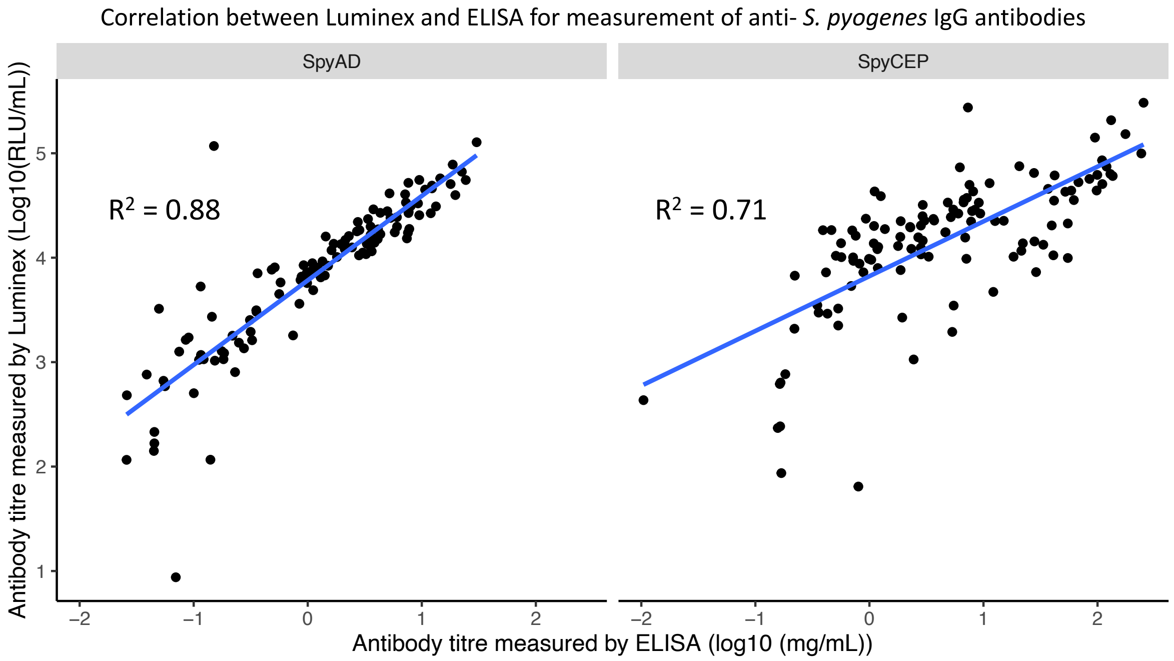


**Figure S2:** Correlation between IgG antibody titres measured from individual participants using ELISA and Luminex platforms. Log10 transformed IVIG-adjusted anti- protein activity for ELISA and Relative Luminex Units (RLU) for Luminex were analysed for correlation with Pearson method. Antigens used in the Luminex assay were obtained from National Institute of Biological Standards and Control, UK. Antigens in Luminex assay were obtained from GSK Vaccine institute for Global Health.

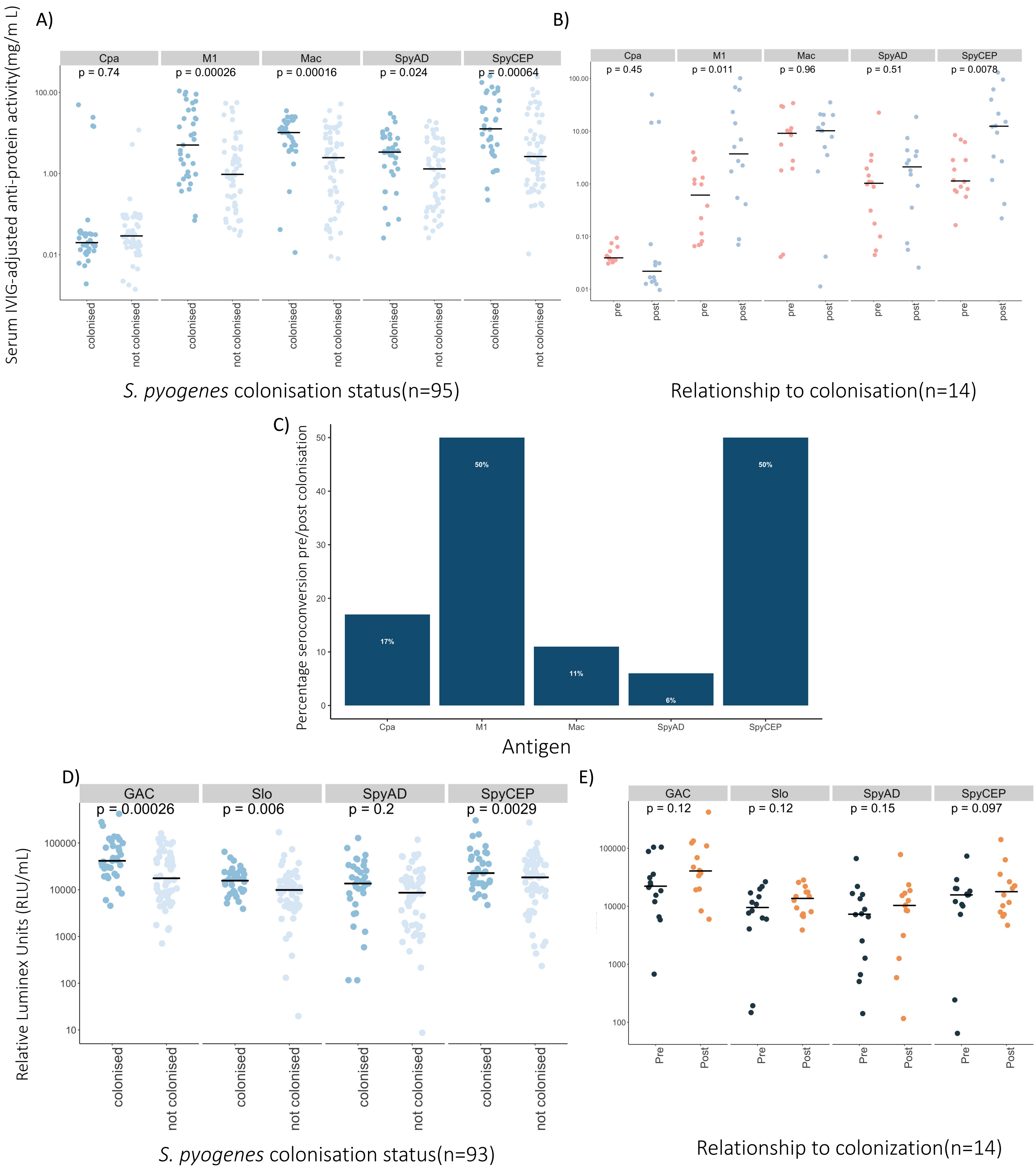

**Figure S3. Serological responses to S. pyogenes colonization measured by ELISA and Luminex 4-plex, excluding 6 participants with sore throat (n=2) and/or infected skin sores (n=5) to ensure symptomatic pharyngitis or skin infection was not driving the serological responses observed in colonized children. A.** Comparison of anti-protein IgG activity to Cpa, M1, Mac, SpyCEP, and SpyAD in participants (n=95) according to anytime S. pyogenes colonization status. **B.** Paired comparison of anti-protein IgG activity to Cpa, M1, Mac, SpyCEP, and SpyAD between day 0 and day 21 in newly colonized participants (n=14). Log10 transformed IVIG-adjusted anti-protein activity was compared with t-tests (unpaired and paired respectively), horizontal line depicts the median value. **C**. Percentage of study participants (n=14) acquiring S. pyogenes during the study who seroconverted between day 0 and day 21. **D.** Comparison of IgG activity to GAC, SLO, SpyCEP, and SpyAD in participants (n=93) according to anytime S. pyogenes colonization status. **E**. Paired comparison of IgG activity to GAC, SLO, SpyCEP, and SpyAD between D0 and D21 in newly-colonized participants (n=14). $Log_{10}$ transformed IVIG-adjusted anti-antigen activity was compared with t-tests (unpaired and paired respectively). Horizontal bar depicts the median value.

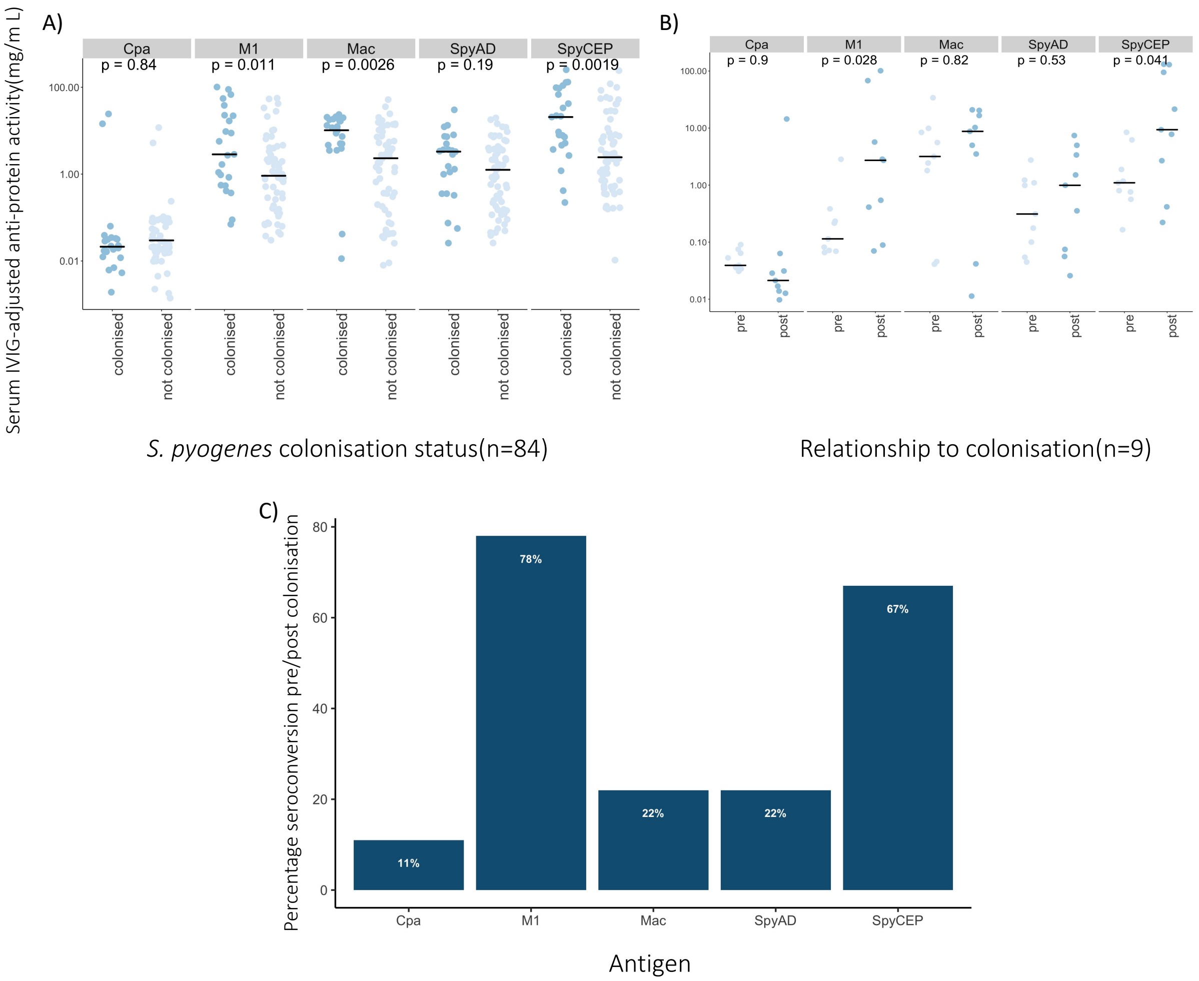


**Figure S4. Serological responses to S. pyogenes colonization measured by ELISA excluding participants who were only colonized at D21, thereby attributing any serological responses to events occurring at least 14 days prior to serological measurement. A.** Comparison of anti-protein IgG activity to Cpa, M1, Mac, SpyCEP, and SpyAD in participants (n=84) according to S. pyogenes colonization status on D0 or D7 of study. **B**. Paired comparison of anti-protein IgG activity to Cpa, M1, Mac, SpyCEP, and SpyAD between day 0 and day 21 in participants newly colonized (n=9) on D7 of study. $Log_{10}$ transformed IVIG-adjusted anti-protein activity was compared with t-tests (unpaired and paired respectively), horizontal line depicts the median value. **C**. Percentage of study participants (n=18) acquiring S. pyogenes during the study who seroconverted between day 0 and day 21.

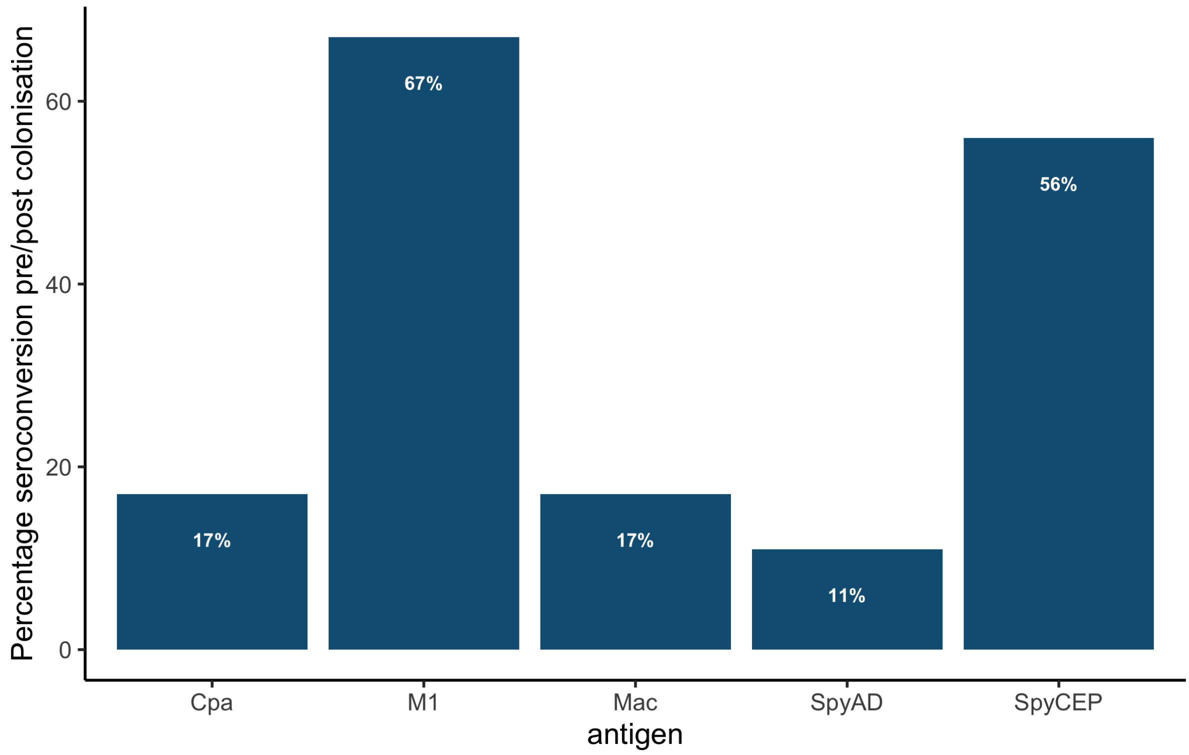


**Figure S5: Percentage of newly-colonized study participants (n=18) who seroconverted between day 0 and day 21, using a more conservative definition of 4-fold increase in IgG titre measured by ELISA.**
